# Efficacy and Safety of Sodium-Glucose Cotransporter 2 Inhibitors in the Very Elderly with Chronic Kidney Disease: a Retrospective Cohort Study Using Real-World Data

**DOI:** 10.64898/2026.08.11.26360158

**Authors:** Jonas Michael Willerding, Anette Melk, Kai Schmidt-Ott, Robert Greite, Wilfried Gwinner, Julian Doricic, Bernhard MW Schmidt

**Affiliations:** Department of Nephrology and Hypertension, Hannover Medical School, Hannover, Germany; Department of Pediatric Kidney, Liver and Metabolic Diseases and Neuropediatrics, Hannover Medical School, Hannover, Germany

**Keywords:** CKD, SGLT2 inhibitors, sodium-glucose cotransporter 2 inhibitors elderly, 80 years

## Abstract

**Importance:** The prevalence of chronic kidney disease (CKD) is increasing, with aging being a major contributor. Sodium-glucose cotransporter 2 inhibitors (SGLT2i) are well established therapies for CKD; however, adults aged 80 years or older have been underrepresented in previous large-scale trials. Consequently, evidence regarding effects of SGLT2i therapy in this cohort remains limited.

**Objective:** To evaluate the association of SGLT2i initiation with mortality, kidney outcomes and cardiovascular outcomes among patients over 80 years of age and CKD.

**Design, Setting, Participants:** This retrospective cohort study used data from TriNetX Research Network, a multicenter electronic health record database. To ensure comparable standard of care and SGLT2i eligibility, the period for the occurrence of the index event was restricted to January 1, 2021, until January 1, 2025. Propensity score matching was performed to balance comorbidities, laboratory parameters, concomitant medications, and frailty-associated factors between groups.

**Exposures:** Initiation of SGLT2i therapy vs. non-use

**Main Outcomes and Measures:** Outcome analysis focused on all-cause mortality, major adverse kidney events (MAKE) and major adverse cardiovascular events (MACE). Cox proportional hazards models were used to estimate hazard ratios with 95% confidence intervals; following propensity score matching, results were considered as adjusted hazard ratios (aHR).

**Results:** After propensity score matching, 5,038 patients were included in each group. During two years of follow-up, SGLT2i initiation was associated with lower all-cause mortality (aHR 0.818; 95% CI 0.746 – 0.896, p<0.0001) and fewer MAKE events (aHR 0.779; 95% CI 0.0.715 - 0.850, p<0.0001). No significant difference in MACE was observed (aHR 1.007; 95%CI 0.938 - 1.082, p=0.84). Results were generally consistent across subgroups. Risk of acute kidney injury was higher in the SGLT2i group, while incident dialysis and end-stage renal disease were significantly reduced. Acute myocardial infarction and acute heart failure were increased in the SGLT2i group.

**Conclusion and Relevance:** Regarding survival and kidney endpoints, even the oldest CKD patients seem to benefit from SGLT2i treatment, which was associated with reduced mortality as well as improved long-term renal outcomes. MACE showed no significant differences, while specific cardiac events were increased, reflecting possible safety concerns requiring further investigation in this specific age group.

**Key points:** *Question:* Is sodium-glucose cotransporter 2 inhibitor (SGLT2i) initiation associated with clinical outcomes in adults aged 80 years or older with chronic kidney disease?

*Findings:* In this retrospective cohort study of 10,076 propensity-score matched adults aged 80 years or older with chronic kidney disease, SGLT2i initiation was associated with lower all-cause mortality and fewer major adverse kidney events over 2 years.

*Meanings:* These findings support the consideration of SGLT2i treatment in very old adults with chronic kidney disease, a population underrepresented in clinical trials.

## INTRODUCTION

The global burden of chronic kidney disease (CKD) grows continuously and population aging has been identified as major driver. Notably, CKD incidence, prevalence and mortality increases with age, with highest incidence in individuals aged between 80 to 84 years.^1,2^ Demographic shifts are expected to further increase the number of patients with CKD.^3–6^ Managing CKD in the elderly is essential, as advanced age is linked to increased cardiovascular risk and mortality which is further amplified by CKD. Effective care may reduce complications and lead to better quality of life, especially by preventing end-stage renal disease which would rise questions of whether or not to start kidney replacement therapy in old and frail individuals.

The Kidney Disease: Improving Global Outcomes (KDIGO) CKD guideline underlines the importance of early treatment to slow down progression and reduce cardiovascular risk.^7^ Sodium-glucose cotransporter 2 inhibitors (SGLT2i) have become well established as a treatment for patients with CKD, particularly in patients with albuminuria or concomitant diseases like heart failure or diabetes mellitus. Robust evidence of renoprotective effects and reduced cardiovascular risk was provided by several randomized controlled trials, including CREDENCE^8^, DAPA-CKD^9^, and EMPA-KIDNEY^10^. These findings have since revolutionized CKD treatment.

However, the applicability of this data to elderly patients remains uncertain, since the mean age in the CREDENCE, DAPA-CKD and EMPA-KIDNEY trials was between 62 to 64 years. Subgroup and post hoc analyses for different age groups suggest that elderly patients might experience similar benefits. However, these analyses are limited by small sample size and are not powered to detect efficacy regarding renal and cardiovascular endpoints including all-cause mortality in CKD patients aged over 80 years.^11^ This lack of evidence leads to uncertainty in care providers who are responsible for risk-benefit assessment among older adults. Increased mortality and polypharmacy in this population may diminish the clinical long-term benefits observed in clinical trials. Assumed higher incidence of side effects in older patients – such as urinary tract infection and fractures – influence treatment decisions. Emphasizing this, despite existing recommendations for SGLT2i treatment and patient eligibility, prescription rates within older patient populations remain lower than in other age groups.^12,13^

The underrepresentation of older patients in clinical trials, especially the population over 80 years, results in a gap between guideline recommendations and real-world applicability. Dedicated analyses are required to close this gap. This study evaluated the effect of SGLT2i initiation on mortality and renal outcomes in a large cohort of elderly patients over 80 years of age with CKD using real-world data.

## METHODS

We performed a retrospective cohort study using real-world data from the TriNetX platform (TriNetX LLC, Cambridge, MA, USA).^14^ TriNetX is a global health research network comprising anonymized electronic health records from over 170 healthcare organizations. Data are deidentified and include diagnoses according to the International Classification of Diseases (10^th^ Revision, Clinical Modification, ICD-10-CM), medications according to the U.S. Department of Veteran’s Affairs (VA), the Anatomical Therapeutic Chemical (ATC) or RxNorm classification as well as laboratory values. Codes used for cohort construction, propensity score matching and outcome definitions are presented in Supplementary Table S1. Institutional review board approval and informed consent were waived because all data available through TriNetX are aggregated and deidentified.

### Patient selection and Propensity Score Matching

This study compared initiation of SGLT2i use with no SGLT2i use in patients aged 80 years and beyond with CKD. Cohorts were constructed utilizing the TriNetX Query Builder. Supplementary Figure S1 highlights the study design. Patient selection criteria are listed in Supplementary Tables S2 and S3. General exclusion criteria included type 1 diabetes mellitus and/or a history of kidney transplantation. Patients with an estimated glomerular filtration rate (eGFR) of 20 mL/min/1.73m^2^ or less, requiring renal replacement therapy, or diagnosed with ESRD prior to the index event, were also excluded. The index event was defined as the diagnosis of CKD at ≥ 80 years. For the SGLT2i group, SGLT2i had to be initiated within 7 days after the index event and prior use was excluded. In the control group, SGLT2i use was generally excluded. To ensure that all patients received a comparable standard of care and were eligible for SGLT2i treatment for CKD, the period for the occurrence of the index event was restricted to January 1^st^, 2021, until January 1^st^, 2025.

For one-to-one propensity score matching (PSM), we used the built-in tool from TriNetX which is based on logistic regression and a greedy nearest-neighbor algorithm with a caliper of 0.1 pooled standard deviation. Parameters including diagnoses, frailty-associated factors, co-medication, and laboratory data were included in the matching process.

### Outcomes

All-cause mortality was the primary outcome. Secondary endpoints included major adverse kidney events (MAKE), defined as a composite of death, dialysis, and end-stage kidney failure and major adverse cardiovascular events (MACE), defined as a composite of death, cardiac arrest, myocardial infarction and cerebral infarction including non-traumatic intracerebral hemorrhage. Individual outcomes in these composites were also analyzed. Individual kidney and cardiovascular events were evaluated. Urinary tract infections (UTI), urogenital candidiasis, and ketoacidosis were investigated as adverse effects. As unrelated outcomes, hematologic neoplasms and gastritis have been examined. Outcome definitions are summarized in Supplementary Table S5. Follow-up ranged from 7 days to 2 years after the index event. Missing data were not filled by statistical imputation. Analyses were based on the available electronic health record data provided by the TriNetX platform. The extent of missing data for individual variables could not be comprehensively assessed.

### Subgroups and sensitivity analyses

Age- and sex-specific subgroups were constructed using the TriNetX platform’s Query Builder and built-in filter function. For cohorts with heart failure and type 2 diabetes mellitus, the respective diagnosis had to be present at or before the index event. For BMI cohorts, patients must have had the respective BMI at least once in the two years prior to or on the index date and must not have had a BMI not according to the threshold in this time period. CKD stage subgroups required a corresponding diagnosis within 2 years before or at the index date. Diagnosis of a more severe CKD stage during the same period was an exclusion criterion. Cohorts of SGLT2i users and non-users were built for every subgroup. To ensure robustness of the primary results, we conducted landmark analyses with the follow-up period starting at predefined time points 3 and 6 months after the index event and spanning until the two-year mark after the index event. Further sensitivity analyses used follow-up periods ending at 3 months until 3 years after the index event.

### Statistics

Cox proportional hazards models were used to estimate hazard ratios with 95% confidence intervals and results were considered as adjusted hazard ratios (aHR), as previous PSM had been performed. Log-rank tests were used to compare the cohorts, with a two-sided p-value of less than 0.05 considered statistically significant. Forest plots were generated in R (version 4.5.2, 2025-10-31, R Foundation for statistical computing, Vienna, Austria).^15^

## RESULTS

### Patient selection and characteristics

Figure 1 illustrates the patient selection process. A total of 5,045 patients receiving SGLT2i and 370,577 patients without SGLT2i treatment were identified. In Table 1 and Supplementary Table S4, patient characteristics before and after PSM are listed. Prior to PSM, patients receiving SGLT2i had a greater burden of cardiovascular comorbidities compared to those without SGLT2i therapy, e.g. presenting higher rates of diabetes mellitus, heart failure, hypertension, and ischemic heart disease. Patients treated with SGLT2i were more likely to receive renin–angiotensin– aldosterone system inhibition and GLP-1 receptor agonist therapy. Following PSM, n=5,038 patients were included in both patient groups, with well-balanced baseline characteristics in terms of age, comorbidities, co-medication, frailty-related factors and laboratory parameters.

**Figure 1:**
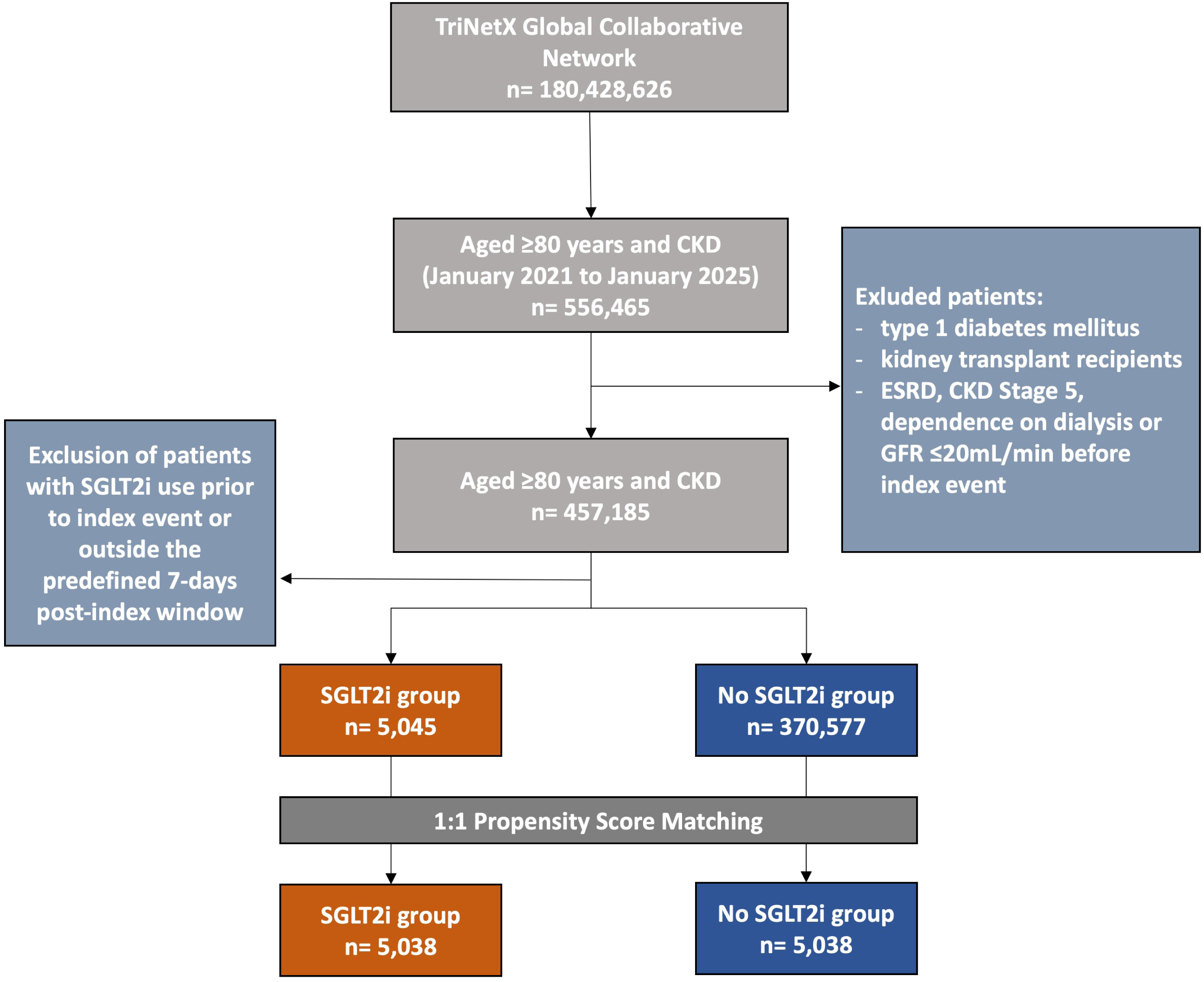
Flowchart of patient selection and PSM process.

**Table 1:** Baseline patient characteristics before and after propensity score matching.

|  | Before matching |  |  | After matching |  |  |
| --- | --- | --- | --- | --- | --- | --- |
|  | SGLT2i<br>users<br>(n= 5,045) | Non-users<br>(n= 370,577) | SMD | SGLT2i<br>users<br>(n= 5,038) | Non-users<br>(n= 5,038) | SMD |
| **Demographics** |  |  |  |  |  |  |
| Age at index,<br>mean ± SD* | 83.0 ± 2.3 | 82.6 ± 2.3 | 0.195 | 83.0 ± 2.3 | 83.0 ± 2.4 | 0.022 |
| Male | 2,959 (58.7%) | 172,793 (46.6%) | 0.243 | 2,952 (58.6%) | 2,982 (59.2%) | 0.012 |
| Female* | 2,079 (41.2%) | 197,434 (53.3%) | 0.244 | 2,079 (41.3%) | 2,050 (40.7%) | 0.012 |
| **Comorbidities, n (%)** |  |  |  |  |  |  |
| Hypertensive<br>diseases* | 4,420 (87.6%) | 304,893 (82.3%) | 0.150 | 4,414 (87.6%) | 4,452 (88.4%) | 0.023 |
| Diabetes mellitus* | 3,341 (66.2%) | 127,695 (34.5%) | 0.670 | 3,334 (66.2%) | 3,429 (68.1%) | 0.040 |
| Overweight and<br>obesity* | 876 (17.4%) | 55,649 (15.0%) | 0.064 | 876 (17.4%) | 943 (18.7%) | 0.035 |
| Disorders of<br>lipoprotein<br>metabolism and other<br>lipidemias* | 3,468 (68.7%) | 229,406 (61.9%) | 0.144 | 3,461 (68.7%) | 3,535 (70.2%) | 0.032 |
| Ischemic heart<br>diseases* | 2,777 (55.0%) | 126,991 (34.3%) | 0.427 | 2,772 (55.0%) | 2,847 (56.5%) | 0.030 |
| Heart failure* | 2,983 (59.1%) | 89,010 (24.0%) | 0.762 | 2,977 (59.1%) | 2,983 (59.2%) | 0.002 |
| Cardiomyopathy* | 764 (15.1%) | 18,618 (5.0%) | 0.341 | 760 (15.1%) | 778 (15.4%) | 0.010 |
| Atrial fibrillation and<br>flutter | 2,104 (41.7%) | 99,445 (26.8%) | 0.317 | 2,100 (41.7%) | 2,070 (41.1%) | 0.012 |
| Cerebrovascular<br>diseases* | 838 (16.6%) | 63,198 (17.1%) | 0.012 | 836 (16.6%) | 862 (17.1%) | 0.014 |
| Atherosclerosis* | 456 (9.0%) | 33,746 (9.1%) | 0.002 | 455 (9.0%) | 436 (8.7%) | 0.013 |
| Chronic kidney<br>disease, stage 1 | 45 (0.9%) | 4,509 (1.2%) | 0.032 | 44 (0.9%) | 84 (1.7%) | 0.071 |
| Chronic kidney<br>disease, stage 2* | 362 (7.2%) | 33,540 (9.1%) | 0.069 | 362 (7.2%) | 362 (7.2%) | <0.001 |
| Chronic kidney<br>disease, stage 3* | 3,056 (60.6%) | 242,438 (65.4%) | 0.101 | 3,054 (60.6%) | 3,056 (60.7%) | 0.001 |
| Chronic kidney<br>disease, stage 4* | 353 (7.0%) | 30,244<br>(8.2%) | 0.044 | 352 (7.0%) | 359 (7.1%) | 0.005 |
| Acute kidney failure* | 1,737 (34.4%) | 75,684 (20.4%) | 0.318 | 1,734 (34.4%) | 1,711 (34.0%) | 0.010 |
| Neoplasms* | 1,029 (20.4%) | 108,402 (29.3%) | 0.206 | 1,029 (20.4%) | 1,042 (20.7%) | 0.006 |
| Aplastic and other<br>anemias and other<br>bone marrow failure<br>syndromes* | 1,443 (28.6%) | 103,184 (27.8%) | 0.017 | 1,440 (28.6%) | 1,443 (28.6%) | 0.001 |
| Candidiasis of vulva<br>and vagina* | 12 (0.2%) | 1,361 (0.3%) | 0.024 | 12 (0.2%) | 12 (0.2%) | <0.001 |
| Candidiasis of other<br>urogenital sites* | 21 (0.4%) | 785 (0.2%) | 0.037 | 21 (0.4%) | 19 (0.4%) | 0.006 |
| Urinary tract infection,<br>site not specified* | 509 (10.1%) | 52,995 (14.3%) | 0.129 | 509 (10.1%) | 544 (10.8%) | 0.023 |
| Cystitis* | 188 (3.7%) | 25,637 (6.9%) | 0.143 | 188 (3.7%) | 191 (3.8%) | 0.003 |
| Acute pyelonephritis | 11 (0.2%) | 1,562 (0.4%) | 0.036 | 11 (0.2%) | 12 (0.2%) | 0.004 |
| Slipping, tripping, stumbling and falls* | 541 (10.7%) | 45,397 (12.3%) | 0.048 | 541 (10.7%) | 561 (11.1%) | 0.013 |
| Problems related to care provider dependency* | 286 (5.7%) | 14,214 (3.8%) | 0.086 | 286 (5.7%) | 294 (5.8%) | 0.007 |
| Unspecified dementia* | 370 (7.3%) | 31,932 (8.6%) | 0.047 | 370 (7.3%) | 379 (7.5%) | 0.007 |
| Vascular dementia* | 64 (1.3%) | 7,618 (2.1%) | 0.062 | 64 (1.3%) | 71 (1.4%) | 0.012 |
| **Medication, n (%)** |  |  |  |  |  |  |
| ACE-Inhibitors* | 797 (15.8%) | 53,642 (14.5%) | 0.037 | 795 (15.8%) | 839 (16.7%) | 0.024 |
| Angiotensin II Inhibitors* | 1,560 (30.9%) | 70,555 (19.0%) | 0.277 | 1,554 (30.8%) | 1,549 (30.7%) | 0.002 |
| GLP-1 analogues* | 193 (3.8%) | 4,441 (1.2%) | 0.168 | 191 (3.8%) | 196 (3.9%) | 0.005 |
| Beta-Blockers* | 2,467 (48.9%) | 124,653 (33.6%) | 0.314 | 2,463 (48.9%) | 2,539 (50.4%) | 0.030 |
| Finerenone* | 21 (0.4%) | 111 (0.0%) | 0.082 | 19 (0.4%) | 12 (0.2%) | 0.025 |
| Spironolactone* | 538 (10.7%) | 15,560 (4.2%) | 0.248 | 536 (10.6%) | 525 (10.4%) | 0.007 |
| Eplerenone* | 48 (1.0%) | 741 (0.2%) | 0.099 | 48 (1.0%) | 37 (0.7%) | 0.024 |
| Loop Diuretics* | 2,206 (43.7%) | 72,297 (19.5%) | 0.539 | 2,201 (43.7%) | 2,166 (43.0%) | 0.014 |
| Thiazides* | 685 (13.6%) | 49,669 (13.4%) | 0.005 | 684 (13.6%) | 711 (14.1%) | 0.016 |
| Metformin* | 842 (16.7%) | 24,941 (6.7%) | 0.314 | 837 (16.6%) | 892 (17.7%) | 0.029 |
| Glucocorticoids* | 1,359 (26.9%) | 115,386 (31.1%) | 0.093 | 1,359 (27.0%) | 1,409 (28.0%) | 0.022 |
| Immune suppressants* | 51 (1.0%) | 4,226 (1.1%) | 0.013 | 51 (1.0%) | 48 (1.0%) | 0.006 |
| **Laboratory** |  |  |  |  |  |  |
| eGFR, mL/min/1.73m<sup>2</sup> (CKD-EPI 2021), Mean ± SD | 46.3 ± 14.4 | 48.6 ± 17.0 | 0.150 | 46.3 ± 14.4 | 46.7 ± 16.5 | 0.028 |
| BMI (kg/m<sup>2</sup>), Mean ± SD | 28.1 ± 5.9 | 27.8 ± 5.8 | 0.044 | 28.1 ± 5.9 | 28.3 ± 6.0 | 0.044 |
| HbA1c (%), Mean ± SD | 7.0 ± 1.4 | 6.3 ± 1.1 | 0.541 | 7.0 ± 1.4 | 6.6 ± 1.2 | 0.253 |
| Blood pressure, systolic (mmHg)* | 130.6 ± 22.8 | 133.2 ± 20.9 | 0.119 | 130.6 ± 22.8 | 131.8 ± 22.8 | 0.055 |
| UACR, mg/g Creatinine, Mean ± SD | 528.1 ± 1501 | 482.1 ± 6880 | 0.009 | 532.6 ± 1507 | 195.2 ± 431.3 | 0.304 |
| <30 | 48 (1.0%) | 6,551 (1.8%) | 0.071 | 47 (0.9%) | 99 (2.0%) | 0.086 |
| 30 – 300 | 54 (1.1%) | 5,117 (1.4%) | 0.028 | 54 (1.1%) | 92 (1.8%) | 0.063 |
| 300 – 3000 | 31 (0.6%) | 1,652 (0.4%) | 0.023 | 31 (0.6%) | 36 (0.7%) | 0.012 |
| > 3000 | 10 (0.2%) | 250 (0.1) | 0.036 | 10 (0.2%) | 10 (0.2%) | <0.001 |

**Abbreviations:** ACE, angiotensin-converting-enzyme; BMI, body mass index; eGFR, estimated glomerular filtration rate; HbA1c, glycated hemoglobin; SD, standard deviation; SMD, standardized mean difference; Std. diff., standard difference; UACR, urine albumin-creatinine ratio. More factors at baseline can be found in supplementary table 3.

### Main outcomes

Table 2 shows the outcome comparisons after PSM for mortality, MACE and MAKE between SGLT2i users and non-users. SGLT2i treatment was associated with a significant reduction in all-cause mortality among patients over 80 years of age with CKD. Whereas only 815 deaths occurred in the SGLT2i group, 1,044 patients died in the non-user group during the observation period (aHR 0.818; 95% CI 0.746 - 0.896; p<0.0001). Advantages of SGLT2i use were also observed for MAKE. SGLT2i use was linked to significantly less adverse renal outcomes (aHR 0.779; 95% CI 0.715-0.850; p<0.0001). Regarding MACE, the groups showed no significant difference (aHR 1,007; 95% CI 0.938-1.082; p=0.85).

**Table 2:** Outcome comparison between SGLT2i users and no-SGLT2i users.

| Outcomes | No. of patients with outcome |  | aHR | 95% CI | p |
| --- | --- | --- | --- | --- | --- |
|  | SGLT2i (n=5,038) | no-SGLT2i (n=5,038) |  |  |  |
| All-cause mortality | 815 | 1,044 | 0.818 | 0.746 – 0.896 | <0.0001 |
| MAKE | 902 | 1,194 | 0.779 | 0.715 – 0.850 | <0.0001 |
| MACE | 1,459 | 1,548 | 1.007 | 0.938 – 1.082 | 0.8413 |

Abbreviations: aHR, adjusted hazard ratio; CI, confidence interval; MAKE, major adverse kidney events; MACE, major adverse cardiovascular events.

### Subgroup and sensitivity analyses

Outcome comparisons for all-cause mortality and MAKE among investigated subgroups are illustrated in Figure 2 and 3, for MACE in Figure S2. Across most predefined subgroups, SGLT2i treatment was consistently associated with a lower risk of mortality and MAKE. In some subgroups with small patient counts, there was no statistically significant effect although a similar numerical trend was observed. Beneficial effects regarding mortality were more pronounced in patients with type 2 diabetes mellitus (T2DM) (aHR 0.781; 95% CI 0.699 - 0.873). Both survival and kidney benefits were pronounced in patients without heart failure (aHR 0.712; 95% CI 0.588 – 0.862 for mortality; aHR 0.699; 95% CI 0.589 - 0.828 for MAKE) and in those with BMI ≥ 30 (aHR 0.726; 95% CI 0.577 – 0.912 for mortality, aHR 0.750; 95% CI 0.603 - 0.933 for MAKE).

**Figure 2:**
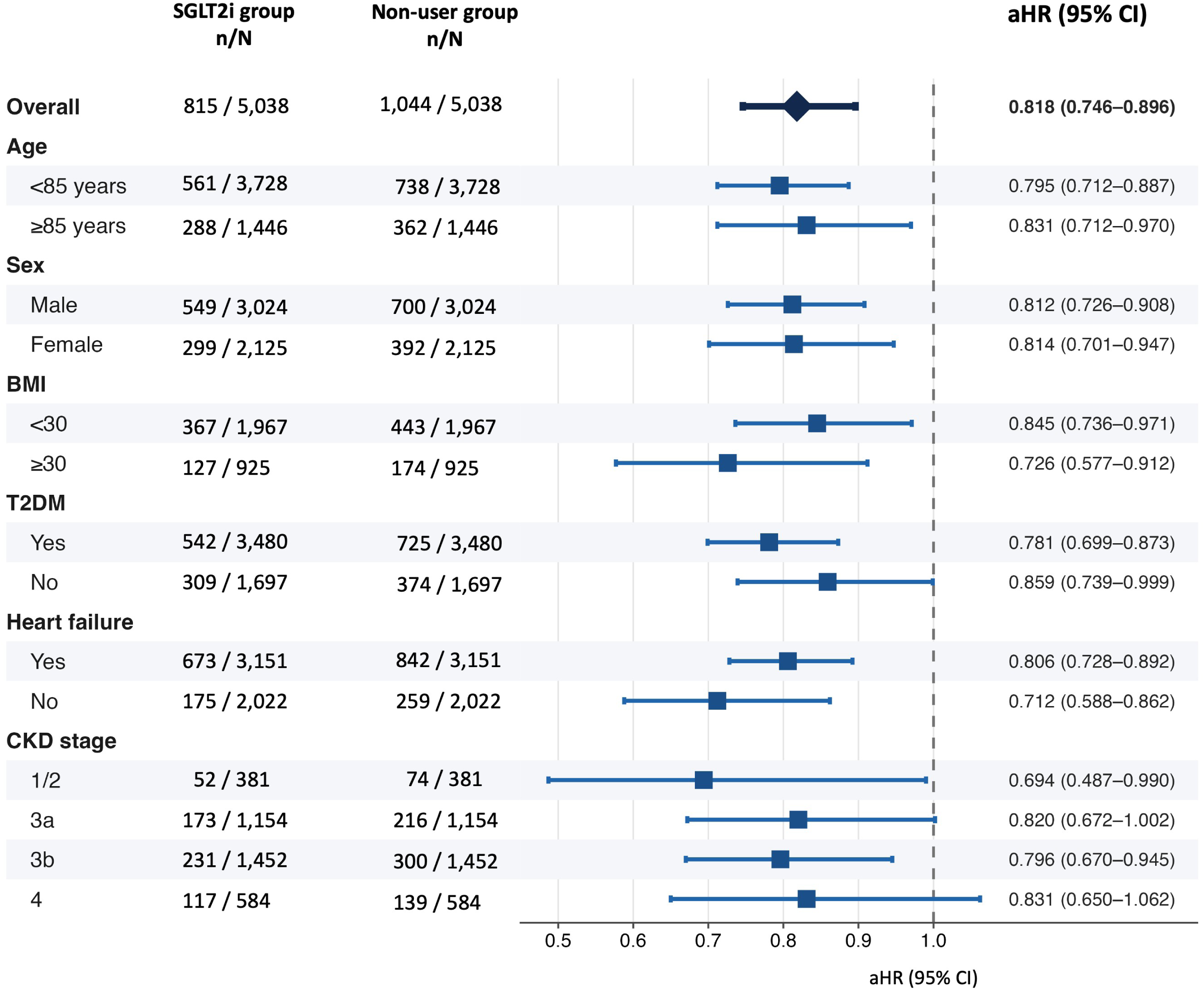
Subgroup analysis for mortality. Forest plots showing subgroup analysis regarding mortality for SGLT2i users versus non-users with adjusted hazard ratios (aHR) and 95% CI. n=events (deaths), N=patients in cohort. Because PSM was repeated for every subgroup investigated, addition of patients in subgroups deviates from the initial cohort size. <u>Abbreviations:</u> BMI, body mass index; T2DM, type 2 diabetes mellitus; CKD, chronic kidney disease; aHR, adjusted hazard ratio; 95% CI, 95% confidence interval

**Figure 3:**
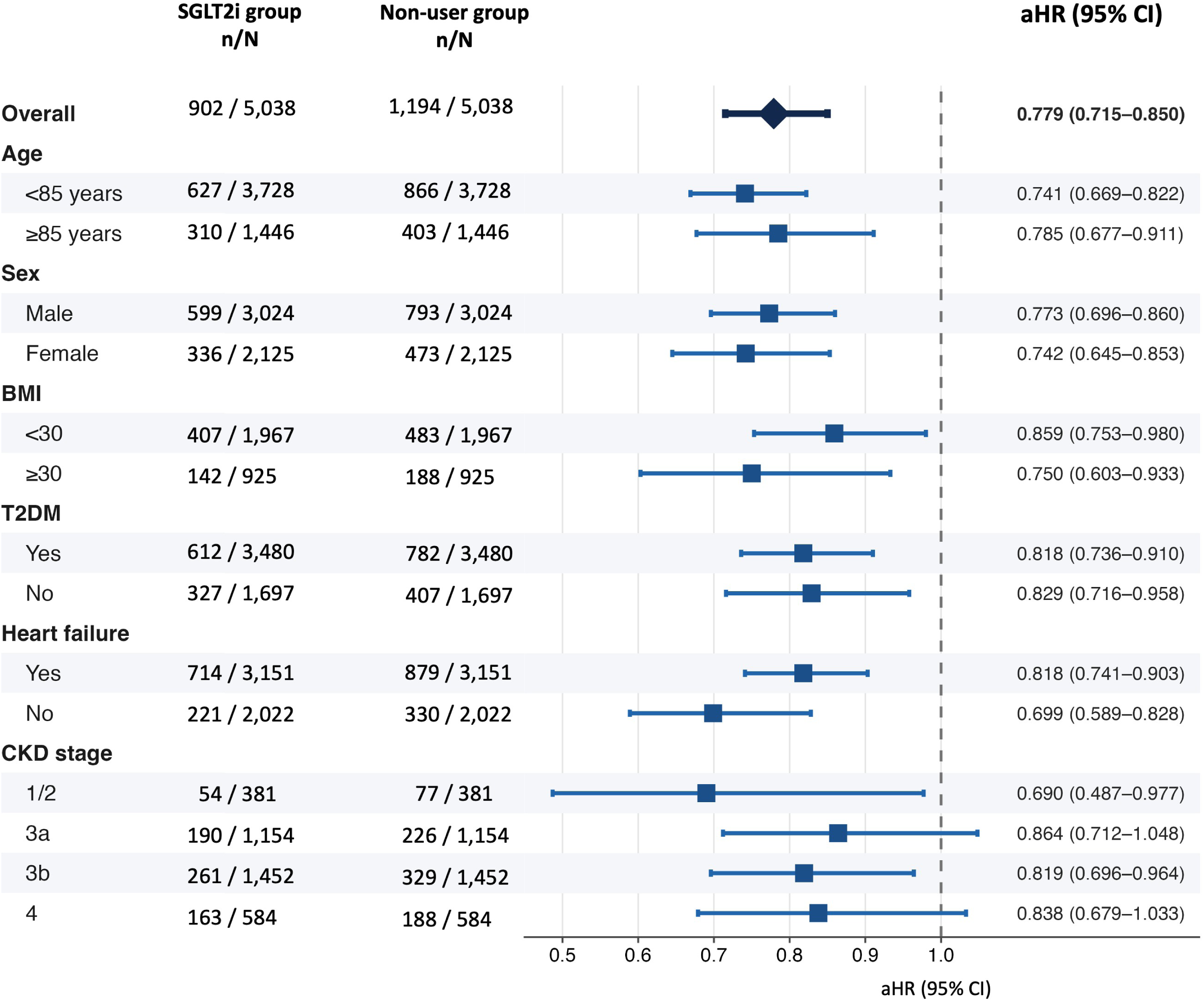
Subgroup analysis for MAKE. Forest plots showing subgroup analysis regarding MAKE for SGLT2i users versus non-users with adjusted hazard ratios (aHR) and 95% CI. n=events, N=patients in cohort. Because PSM was repeated for every subgroup investigated, addition of patients in subgroups deviates from the initial cohort size. <u>Abbreviations:</u> BMI, body mass index; T2DM, type 2 diabetes mellitus; CKD, chronic kidney disease; aHR, adjusted hazard ratio; 95% CI, 95% confidence interval

Across all subgroups, no significant differences in MACE were observed.

Landmark analyses were conducted with the follow-up period starting 3 and 6 months after the index event and spanning until 2 years after index. These analyses confirmed the association of significantly reduced mortality and kidney outcomes with SGLT2i use (Suppl. Table S6). There were no significant differences between cohorts regarding MACE, also supporting the results of our main analysis. Further sensitivity analyses after different follow-up periods showed consistent results at 6, 12 and 18 months as well as 3 years after the index event (Suppl. Table S7). However, with the follow-up period spanning from 7 days to 3 months after the index event, no significant differences in mortality and MAKE were observed. Furthermore, in this follow-up period, the MACE composite was significantly increased.

### Specific renal and cardiovascular events

We observed increased AKI events in the SGLT2i cohort (Fig. 4, aHR 1.107, 95% CI 1.024 - 1.197) while incident dialysis and ESRD were reduced (aHR 0.513, 95% CI 0.377-0.700 for dialysis; aHR 0.569, 95% CI 0.446-0.726 for ESRD). Regarding cardiovascular outcomes, there were no significant differences in the occurrence of cerebral bleeding, cerebral infarction or cardiac arrest (Fig. 4). However, acute myocardial infarction (AMI) and acute heart failure were significantly increased in the SGLT2i cohort (aHR 1.426, 95% CI 1.251-1.625 for AMI; aHR 1.517, 95% CI 1.386-1.661 for acute heart failure).

**Figure 4:**
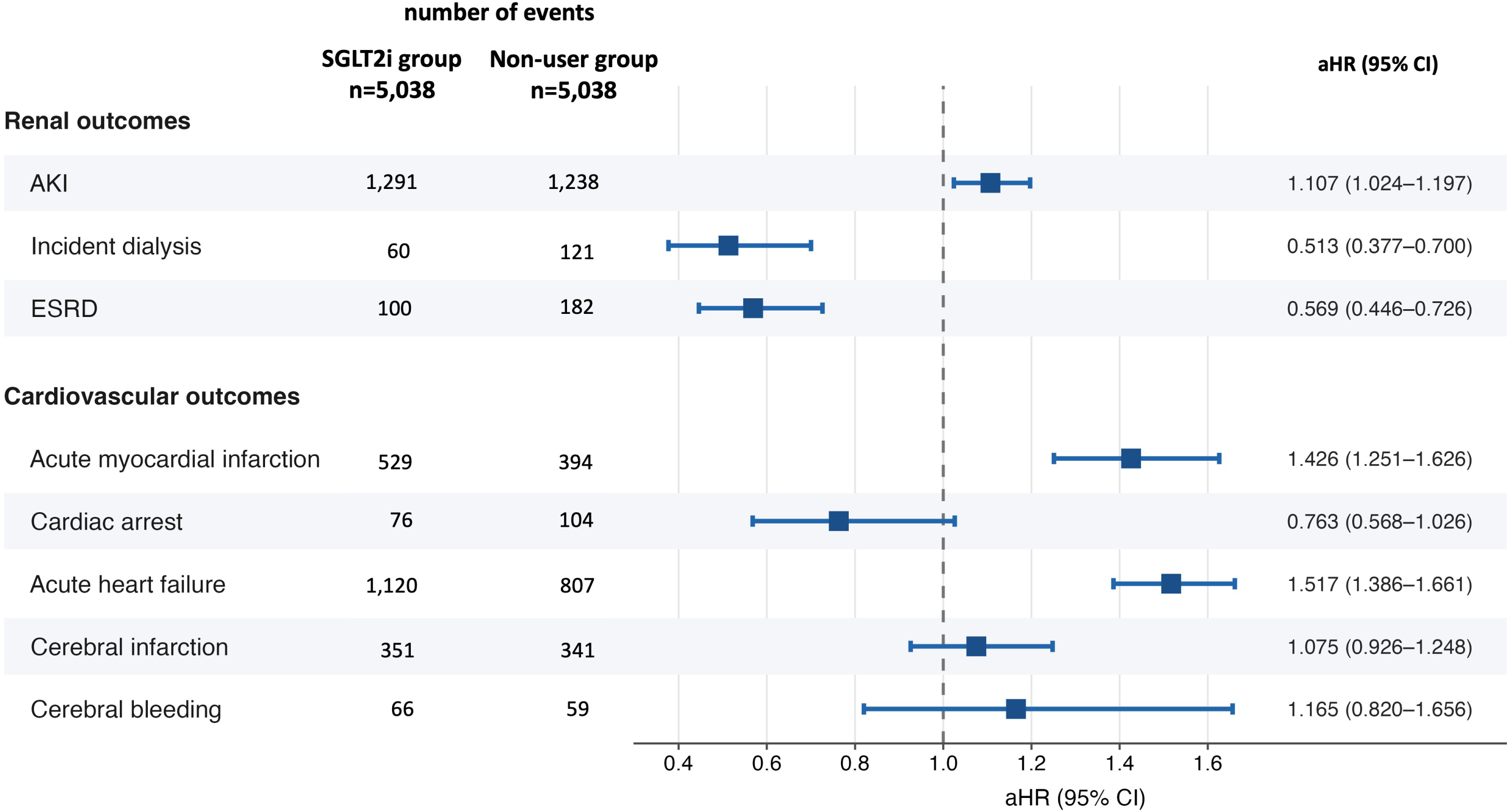
Individual kidney and cardiovascular outcomes. Forest plots for individual kidney and cardiovascular events with adjusted hazard ratios (aHR) and 95% confidence intervals (95% CI). <u>Abbreviations:</u> AKI, acute idney injury; ESRD, end-stage renal disease

### Adverse events and unrelated outcomes

With regard to adverse events, the group treated with SGLT2i did not experience a higher risk of UTI, osteoporotic fractures or ketoacidosis (Fig. 5). As expected, the risk of urogenital candidiasis was significantly higher. In order to support specificity of the observed treatment effects, we examined the occurrence of unrelated outcomes in both cohorts: no differences between the groups were detected concerning hematologic neoplasms and gastritis or duodenitis.

**Figure 5:**
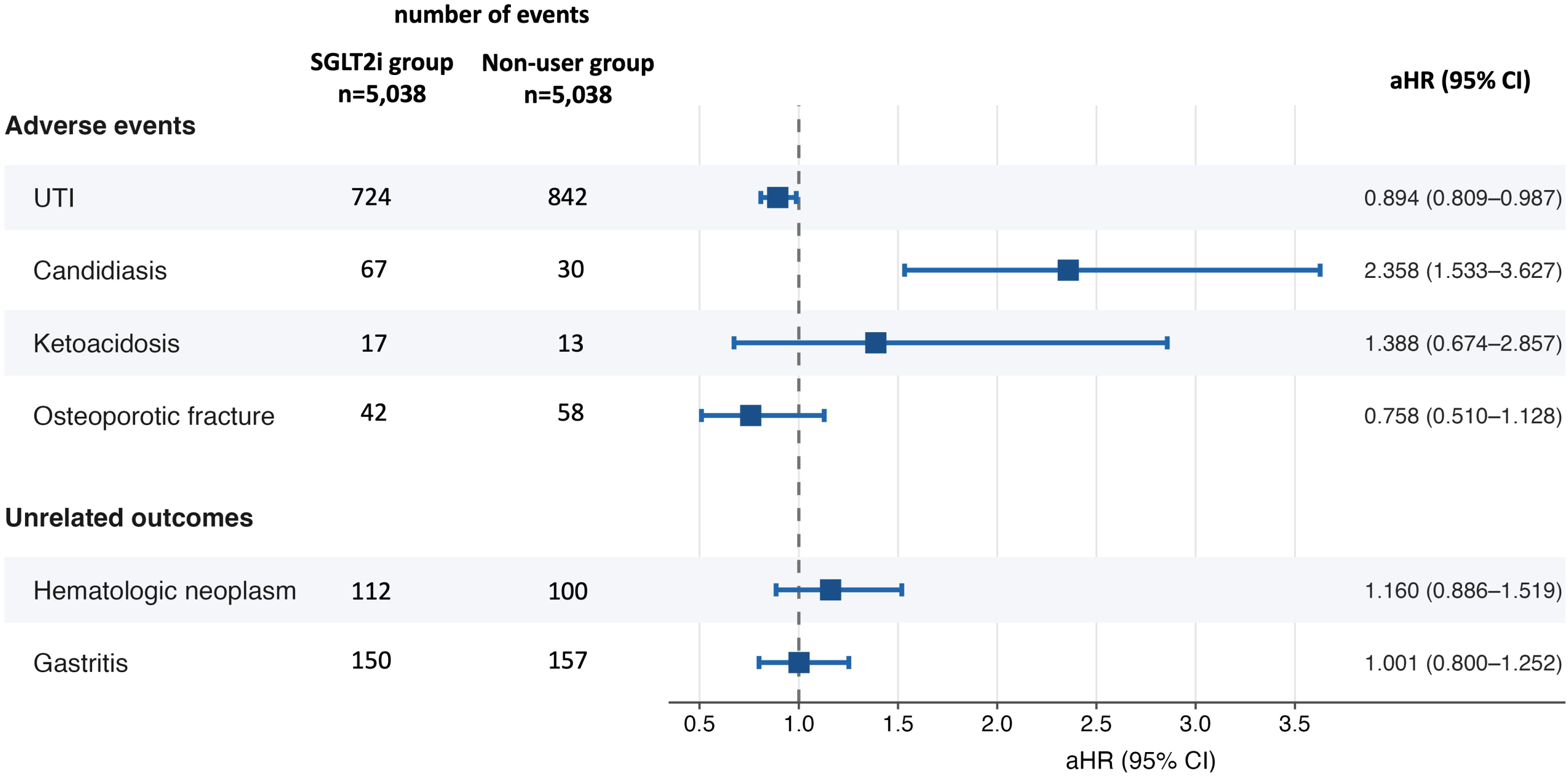
Adverse events and unrelated outcomes. Forest plots for adverse events and unrelated outcomes with adjusted hazard ratios (aHR) and 95% confidence intervals (95% CI). <u>Abbreviations:</u> UTI, urinary tract infections

## DISCUSSION

Our study provides real-world evidence supporting effectiveness of SGLT2i in reduction of mortality as well as major kidney outcomes in patients ≥80 years of age with CKD. However, safety concerns arise from increased AMI and acute heart failure events. These findings help in assessing the risk-benefit relationship in a patient population becoming increasingly relevant in clinical practice. This is particularly important because older adults are and will likely remain underrepresented in randomized controlled trials despite their substantial heterogeneity.

The beneficial effects on survival and MAKE reaffirm trends previously reported in subgroup and post-hoc analyses of large randomized trials. Across the landmark CKD-focused SGLT2i trials (CREDENCE^8^, DAPA-CKD^9^, EMPA-KIDNEY^10^), all-cause mortality was only assessed as secondary outcome, while primary endpoints were composite renal outcomes with or without cardiovascular death. The EMPA-KIDNEY trial with 6,609 participants and mean age of 63.8 years demonstrated the efficacy of empagliflozin in reducing the composite endpoint of kidney disease progression and cardiovascular death in CKD patients. In the subgroup of patients over 70 years, a 35% relative risk reduction for the primary composite endpoint was found (HR 0.65; 95% CI, 0.52 - 0.81). A post-hoc analysis of the CREDENCE trial also analyzed age-specific subgroups.^11^ While the primary composite (kidney failure, doubling of serum creatinine or death due to kidney or cardiovascular disease) was significantly reduced in patients <60 and 60-69, there was only a non-significant trend with weaker effect size in those >70 years (HR 0.89; 95% CI 0.61–1.29; p = 0.5). The DAPA-CKD study provides the most granular data on older individuals.^16^ Septuagenarians and octogenarians represented 25% (1,197 patients) of the study cohort with 198 patients ≥80 years. In the 198 included octogenarians, a non-significant trend in reduction of the primary endpoint was detected (composite of eGFR decline ≥50%, ESRD or death from a kidney- or cardiovascular-related cause; HR 0.72; 95% CI 0.32–1.64; p = 0.440). Regarding frailty factors, a post-hoc analysis demonstrated consistent benefits of dapagliflozin in CKD patients across the frailty spectrum.^17^

Several meta-analyses of randomized controlled trials have demonstrated preserved cardiovascular benefits of SGLT2i in older individuals, including those aged 75 years or older, with some evidence extending to those aged 80 years or older.^18–21^ However, CKD-specific evidence in octogenarians remains limited.

Although SGLT2i were associated with lower mortality and MAKE, we observed increased incidences of AMI and acute heart failure, findings that contrast the overall benefits reported in previous analyses.^18–21^ However, these meta-analyses mostly evaluated composite endpoints, which may obscure differential effects on individual outcome. AMI was not systemically evaluated in most prior meta-analyses. Consistent with our results, Shah et al. reported no significant effect on MACE among patients aged 75 years or older despite reductions in other outcomes.^20^ A recent TriNetX study comparing SGLT2i with DPP4i treatment in patients ≥80 years with diabetes mellitus similarly found reduced mortality but increased heart failure events with SGLT2i, while AMI did not differ significantly between groups.^22^ While these findings warrant further investigation of SGLT2i safety in older individuals, particularly across different follow-up periods, residual confounding by indication in our observational study should be considered. Given the well-established benefits of SGLT2i therapy in heart failure and diabetes, despite inclusion in PSM individuals with more severe forms of these conditions might be more likely to receive SGLT2i therapy. Competing risks may also contribute, as mortality and cardiovascular events reduce the population at risk for subsequent acute heart failure and AMI, which are common events in the elderly.

SGLT2i were associated with an increased incidence of AKI while ESRD and dialysis events were reduced. Therefore, long-term renal protection seems to be preserved in the very elderly. However, they might be more susceptible to the hemodynamic changes in the glomerulus induced by SGLT2i causing the well-known initial “dip” in kidney function.^23^ Furthermore, SGLT2i have been associated with lowering of blood pressure in older patients.^24^ This might lead to more AKI events in this cohort while in other cohorts, AKI events were reduced with SGLT2i.^25^ This might particularly affect elderly patients with CKD as this signal was not seen in comparison to DPP4i in diabetic patients.

Regarding common adverse events, genitourinary infections remain a common concern in prescribing SGLT2i.^26–28^ However, these should be divided into UTI and urogenital candidiasis and investigated separately. The influence of SGLT2i on the occurrence of UTIs is controversially debated. Some studies found increased risk of UTI while others did not.^29–33^ We observed no increased UTI events associated with SGLT2i in our study. However, as repeatedly shown by others, genital mycotic infections were increased with SGLT2i treatment.

The beneficial effects of SGLT2i irrespective of diabetes status have been well established.^9,10,34,35^ In our study, effects were consistent across clinically relevant subgroups. As expected, benefits were more pronounced in patients with T2DM and higher BMI. Surprisingly, in patients without heart failure SGLT2i were associated with larger beneficial effects in our cohort. For mortality and MAKE, all subgroups showed lower event rates, except for CKD stages 3a and 4, in which only non-significant trends toward benefit were observed, possibly due to smaller sample sizes.

## LIMITATIONS

We provide data derived from a large patient cohort addressing a clinically relevant and previously unanswered question. However, certain limitations should be taken into account. Despite extensive PSM including frailty-associated factors, diseases and co-medication, residual confounding such as treatment selection bias cannot be fully excluded in an observational study. In our case of very elderly patients, selection bias could have influenced treatment decisions in opposite directions: Physicians might have preferentially selected “fitter” patients with less perceived risk for adverse effects. However, they might also have preferentially selected patients with more severe conditions because of expected treatment benefits. Another limitation is the small fraction of patients without T2DM or heart failure, both conditions in which SGLT2i treatment is well established and guideline-recommended.^36,37^ Treatment effects therefore cannot be fully distinguished between indications. However, this likely reflects the multimorbid collective of elderly CKD patients. Although quality control measures have been established by the TriNetX platform, our study is reliant on the adequacy and completeness of the provided data which itself is reliant on correct diagnosis and respective coding by physicians and health care organizations. Although matching for frailty-related factors, frailty remains difficult to assess accurately, despite potentially representing a more clinically relevant determinant than chronological age in this patient population. Changes in dosage, treatment interruptions, and treatment discontinuations could not be assessed, as granular prescription-level data is not available via TriNetX. Real-world data cannot replace randomized trials, but they help us gain better insight into specific populations where evidence gaps remain. This is particularly relevant for elderly individuals often underrepresented in randomized controlled trials or excluded due to multimorbidity or frailty. Therefore, our study contributes to clinical applicability of SGLT2i in daily patient care.

## CONCLUSION

In conclusion, while significant survival and renal benefits of SGLT2i treatment seem to be preserved in the very elderly, increased AMI and acute heart failure events raise possible safety concerns. Further studies should evaluate specific risks in this fragile patient population to not miss major advantages due to therapeutic nihilism.

## Data availability statement

The data supporting this study’s findings are available from TriNetX, LLC, but restrictions apply to their availability. The data were accessed under license and cannot be publicly shared. Accredited researchers may obtain access through a data use agreement with TriNetX, LLC, which may involve licensing fees.

## Author contribution

J.M.W. and J.D. participated in research concept, research design, data acquisition, statistical analysis, visualization and drafting of the manuscript. K.S.M., A.M., W.G. and R.G. participated in reviewing and writing the contents. B.M.W.S. participated in research concept, supervision and reviewing and writing the contents.

## Abbreviations

aHR: adjusted hazard ratio
BMI: body mass index
CKD: chronic kidney disease
eGFR: estimated glomerular filtration rate
MACE: major adverse cardiovascular events
MAKE: major adverse kidney events
PSM: propensity score matching
SGLT2i: sodium-glucose cotransporter 2 inhibitors
T2DM: type 2 diabetes mellitus
UTI: urinary tract infection

