## Supplementary data for "Efficacy and Safety of Sodium-Glucose Cotransporter 2 Inhibitors in the Very Elderly with Chronic Kidney Disease: a Retrospective Cohort Study Using Real-World Data"

**Supplementary Figure S1: Graphical illustration of study design**

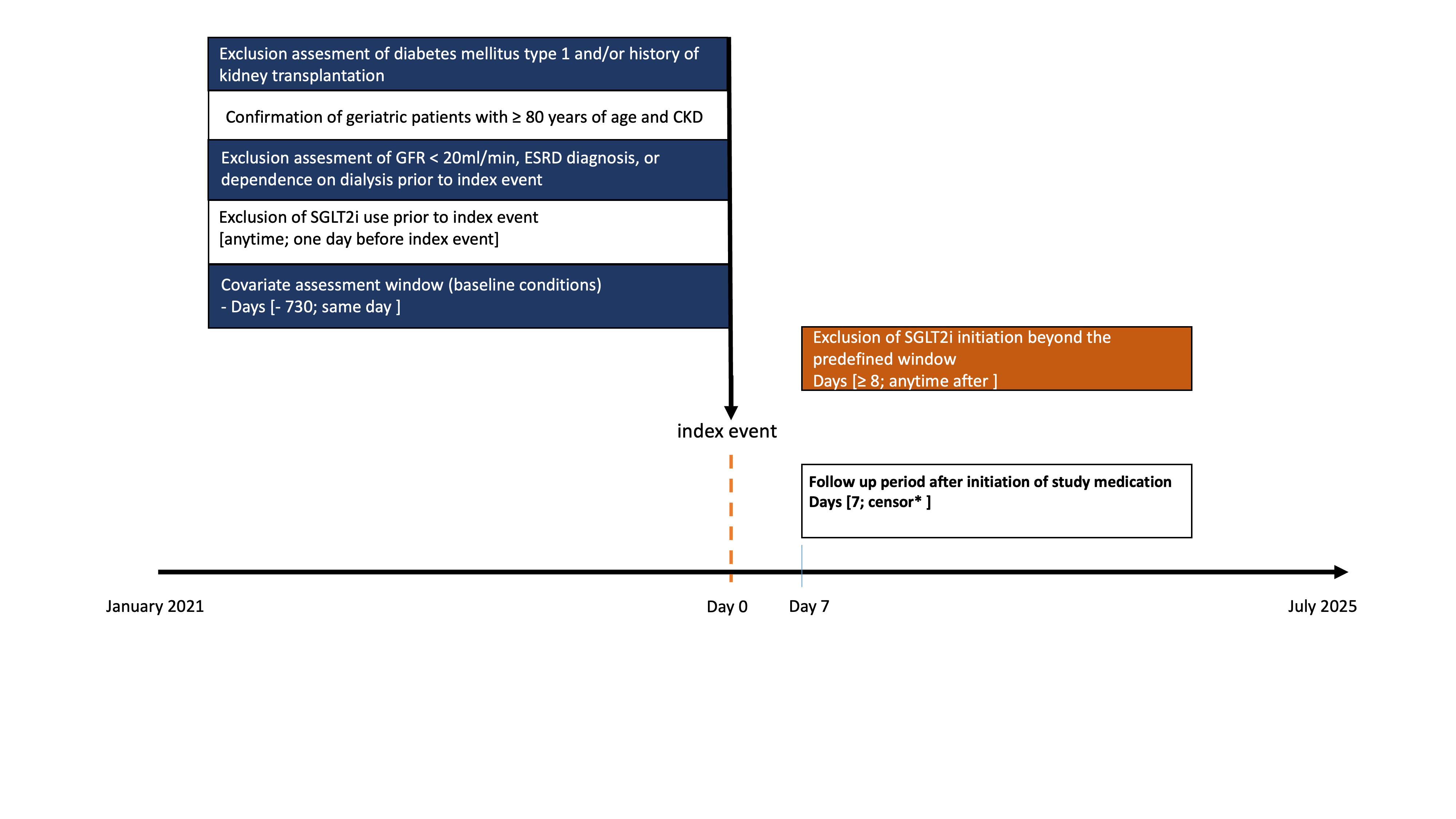

*Occurrence of outcome, death, loss to follow up or end of study period

**Abbreviations**: GFR, glomerular filtration rate; ESRD, end stage renal disease.

**Supplementary Figure S2: Outcome comparison among subgroups for MACE**

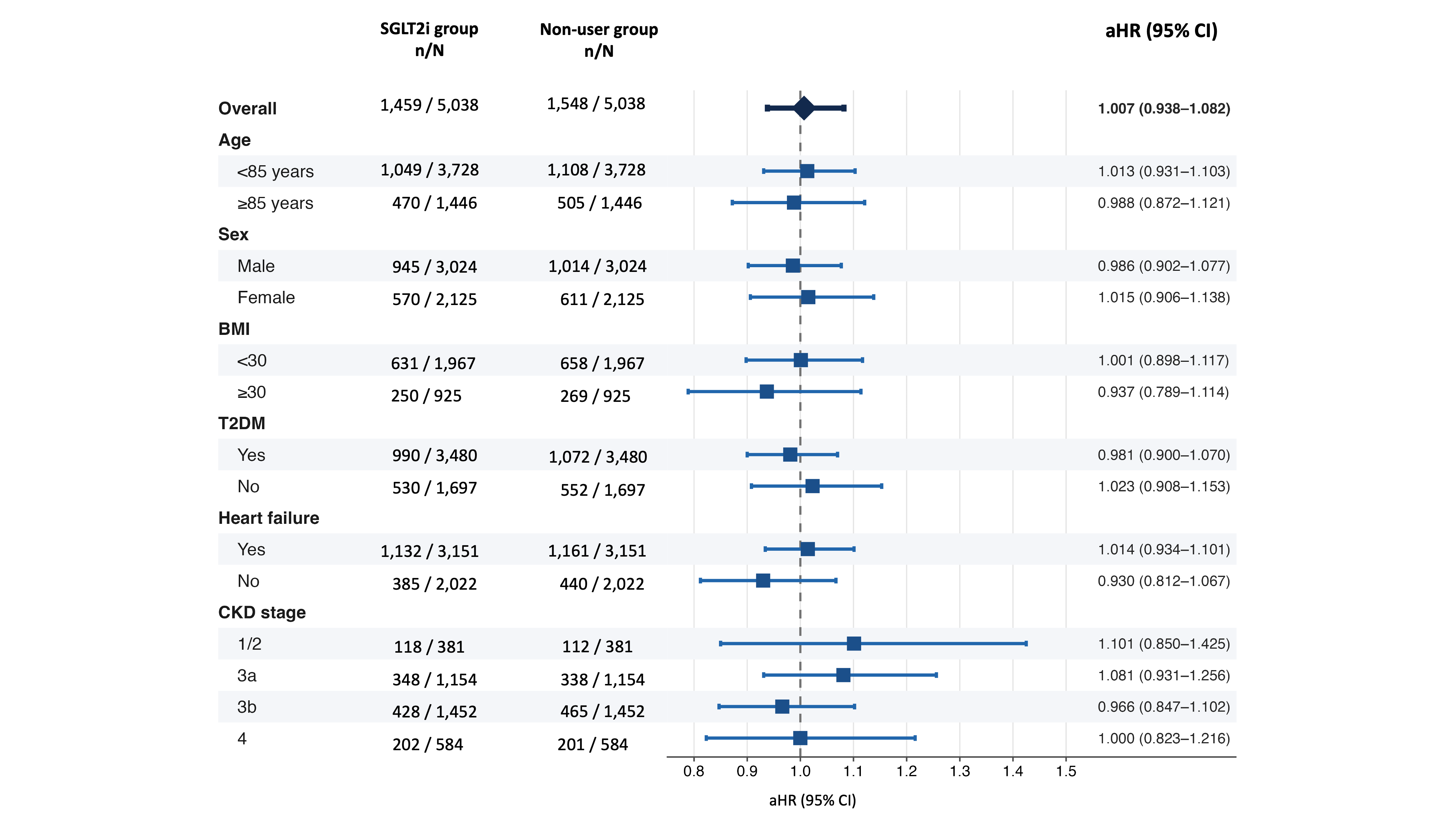

Forest plots showing subgroup analysis regarding MACE for SGLT2i users versus non-users with adjusted hazard ratios (aHR) and 95% CI. n=events, N=patients in cohort. Because PSM was repeated for every subgroup investigated, addition of patients in subgroups deviates from the initial cohort size.

**Abbreviations:** BMI, body mass index; T2DM, type 2 diabetes mellitus; CKD, chronic kidney disease; aHR, adjusted hazard ratio; 95% CI, 95% confidence interval

**Supplementary Table S1: Codes used for Baseline Characteristics / Propensity Score Matching**

| **Demographics** | |
| --- | --- |
| ***Code*** | ***Description*** |
| AI | Age at Index |
| M | Male |
| F | Female |
| UN | Unknown Gender |
| 2054-5 | Black or African American |
| 2028-9 | Asian |
| 2106-3 | White |
| UNK | Unknown Race |
| 2131-1 | Other Race |
| 2186-5 | Not Hispanic or Latino |
| 2135-2 | Hispanic or Latino |
| UN | Unknown Ethnicity |
| **Comorbidities / Diagnoses** | |
| ***Code*** | ***Description*** |
| I10-I1A | Hypertensive diseases |
| E08-E13 | Diabetes mellitus |
| E66 | Overweight and obesity |
| E78 | Disorders of lipoprotein metabolism and other lipidemias |
| I20-I25 | Ischemic heart diseases |
| I05-I09 | Chronic rheumatic heart diseases |
| I26-I28 | Pulmonary heart disease and diseases of pulmonary circulation |
| I48 | Atrial fibrillation and flutter |
| I60-I69 | Cerebrovascular diseases |
| I70 | Atherosclerosis |
| N18.1 | Chronic kidney disease, stage 1 |
| N18.2 | Chronic kidney disease, stage 2 (mild) |
| N18.3 | Chronic kidney disease, stage 3 (moderate) |
| N18.4 | Chronic kidney disease, stage 4 (severe) |
| N18.9 | Chronic kidney disease, unspecified |
| N17 | Acute kidney failure |
| J44 | Other chronic obstructive pulmonary disease |
| F17 | Nicotine dependence |
| K74 | Fibrosis and cirrhosis of liver |
| C00-D49 | Neoplasms |
| D10-D36 | Benign neoplasms, except benign neuroendocrine tumors |
| D37-D48 | Neoplasms of uncertain behavior, polycythemia vera and myelodysplastic syndromes |
| D60-D64 | Aplastic and other anemias and other bone marrow failure syndromes |
| B37.3 | Candidiasis of vulva and vagina |
| B37.4 | Candidiasis of other urogenital sites |
| N39.0 | Urinary tract infection, site not specified |
| N30 | Cystitis |
| N10 | Acute pyelonephritis |
| W00-W19 | Slipping, tripping, stumbling and falls |
| Z74 | Problems related to care provider dependency |
| F03 | Unspecified dementia |
| R06.0 | Dyspnea |
| J96.0 | Acute respiratory failure |
| E87.7 | Fluid overload |
| R07 | Pain in throat and chest |
| I70-I79 | Diseases of arteries, arterioles and capillaries |
| E87.5 | Hyperkalemia |
| E87.6 | Hypokalemia |
| L89 | Pressure ulcer |
| R63.4 | Abnormal weight loss |
| E40-E46 | Malnutrition |
| F01 | Vascular dementia |
| F02 | Dementia in other diseases classified elsewhere |
| Z66-Z66 | Do not resuscitate status (Z66) |
| Z00-Z13 | Persons encountering health services for examinations |
| E87.2 | Acidosis |
| N40 | Benign prostatic hyperplasia |
| I50 | Heart failure |
| I42 | Cardiomyopathy |
| I49 | Other cardiac arrhythmias |
| **Medication** | |
| ***Code*** | ***Description*** |
| CV800 | ACE INHIBITORS |
| CV805 | ANGIOTENSIN II INHIBITOR |
| CV100 | BETA BLOCKERS/RELATED |
| CV050 | DIGITALIS GLYCOSIDES |
| 1656328 | sacubitril |
| 9997 | spironolactone |
| 2562811 | finerenone |
| 298869 | eplerenone |
| CV702 | LOOP DIURETICS |
| CV701 | THIAZIDES/RELATED DIURETICS |
| IM600 | IMMUNE SUPPRESSANTS |
| A10BJ | Glucagon-like peptide-1 (GLP-1) analogues |
| 6809 | Metformin |
| A10BK | Sodium-glucose co-transporter 2 (SGLT2) inhibitors |
| **Laboratory** | |
| ***Code*** | ***Description*** |
| 98979-8 | Glomerular filtration rate/1.73 sq M.predicted [Volume Rate/Area] in Serum, Plasma or Blood by Creatinine-based formula (CKD-EPI 2021) |
| 9083 | BMI |
| 9037 | Hemoglobin A1c/Hemoglobin.total in Blood |
| 9085 | Blood Pressure, Systolic |
| LG34557-5 | Albumin/Creatinine [Mass ratio] in Urine |

**Supplementary Table S2: Query criteria for the SGLT2i cohort**

| Ungrouped terms | | | | | |
| --- | --- | --- | --- | --- | --- |
|  | cannot have |  | diagnosis | UMLS:ICD10CM:E10 | Type 1 diabetes mellitus |
|  |  | or | diagnosis | UMLS:ICD10CM:Z94.0 | Kidney transplant status |
| Group 1 | | | | | |
|  | **Group 1A** | | | | |
|  | must have |  | diagnosis | UMLS:ICD10CM:N18 | Chronic kidney disease (CKD) (at least 80 years old at event) |
|  | date constraint | | The terms in this group occurred between Jan 1, 2021 and Jan 1, 2025 | | |
|  | event relationship | | Any instance of Group 1B occurred on or before the first instance of Group 1A | | |
|  | **Group 1B** | | | | |
|  | cannot have |  | procedure | UMLS:CPT:1012740 | Dialysis Services and Procedures |
|  |  | or | diagnosis | UMLS:ICD10CM:N18.5 | Chronic kidney disease, stage 5 |
|  |  | or | diagnosis | UMLS:ICD10CM:Z99.2 | Dependence on renal dialysis |
|  |  | or | diagnosis | UMLS:ICD10CM:N18.6 | End stage renal disease |
|  |  | or | procedure | UMLS:ICD10PCS:5A1D | Physiological Systems / Performance / Urinary |
|  |  | or | procedure | UMLS:SNOMED:302497006 | Hemodialysis |
|  |  | or | laboratory | UMLS:LNC:62238-1 | Glomerular filtration rate [Volume Rate/Area] in Serum, Plasma or Blood by Creatinine-based formula (CKD-EPI)/1.73 sq M (between 0.00 and 20.00 mL/min/{1.73_m2}) |
|  |  | or | laboratory | TNX:8001 | Glomerular filtration rate/1.73 sq M.predicted [Volume Rate/Area] in Serum, Plasma or Blood by Creatinine-based formula (MDRD) (at most 20.00 mL/min/{1.73_m2}) |
|  |  | or | laboratory | UMLS:LNC:98979-8 | Glomerular filtration rate [Volume Rate/Area] in Serum, Plasma or Blood by Creatinine-based formula (CKD-EPI 2021)/1.73 sq M (at most 20.00 mL/min/{1.73_m2}) |
|  |  | or | laboratory | UMLS:LNC:50210-4 | Glomerular filtration rate [Volume Rate/Area] in Serum, Plasma or Blood by Cystatin C-based formula/1.73 sq M (at most 20.00 mL/min/{1.73_m2}) |
| Group 2 | | | | | |
|  | **Group 2A** | | | | |
|  | must have |  | diagnosis | UMLS:ICD10CM:N18 | Chronic kidney disease (CKD) (at least 80 years old at event) |
|  | date constraint | | The terms in this group occurred between Jan 1, 2021 and Jan 1, 2025 | | |
|  | event relationship | | Any instance of Group 2B occurred at least 1 day before the first instance of Group 2A | | |
|  | **Group 2B** | | | | |
|  | cannot have |  | medication | NLM:ATC:A10BK | Sodium-glucose co-transporter 2 (SGLT2) inhibitors |
| Group 3 | | | | | |
|  | **Group 3A** | | | | |
|  | must have |  | diagnosis | UMLS:ICD10CM:N18 | Chronic kidney disease (CKD) (at least 80 years old at event) |
|  | date constraint | | The terms in this group occurred between Jan 1, 2021 and Jan 1, 2025 | | |
|  | event relationship | | The first instance of Group 3B occurred within 7 days on or after the first instance of Group 3A | | |
|  | **Group 3B** | | | | |
|  | must have |  | medication | NLM:ATC:A10BK | Sodium-glucose co-transporter 2 (SGLT2) inhibitors |

**Supplementary Table S3: Query criteria for the non-SGLT2i cohort**

| Ungrouped terms | | | | | |
| --- | --- | --- | --- | --- | --- |
|  | cannot have |  | diagnosis | UMLS:ICD10CM:E10 | Type 1 diabetes mellitus |
|  |  | or | diagnosis | UMLS:ICD10CM:Z94.0 | Kidney transplant status |
|  |  | or | medication | NLM:ATC:A10BK | Sodium-glucose co-transporter 2 (SGLT2) inhibitors |
| Group 1 | | | | | |
|  | **Group 1A** | | | | |
|  | must have |  | diagnosis | UMLS:ICD10CM:N18 | Chronic kidney disease (CKD) (at least 80 years old at event) |
|  | date constraint | | The terms in this group occurred between Jan 1, 2021 and Jan 1, 2025 | | |
|  | event relationship | | Any instance of Group 1B occurred on or before the first instance of Group 1A | | |
|  | **Group 1B** | | | | |
|  | cannot have |  | procedure | UMLS:CPT:1012740 | Dialysis Services and Procedures |
|  |  | or | diagnosis | UMLS:ICD10CM:N18.5 | Chronic kidney disease, stage 5 |
|  |  | or | diagnosis | UMLS:ICD10CM:Z99.2 | Dependence on renal dialysis |
|  |  | or | diagnosis | UMLS:ICD10CM:N18.6 | End stage renal disease |
|  |  | or | procedure | UMLS:ICD10PCS:5A1D | Physiological Systems / Performance / Urinary |
|  |  | or | procedure | UMLS:SNOMED:302497006 | Hemodialysis |
|  |  | or | laboratory | UMLS:LNC:62238-1 | Glomerular filtration rate [Volume Rate/Area] in Serum, Plasma or Blood by Creatinine-based formula (CKD-EPI)/1.73 sq M (between 0.00 and 20.00 mL/min/{1.73_m2}) |
|  |  | or | laboratory | TNX:8001 | Glomerular filtration rate/1.73 sq M.predicted [Volume Rate/Area] in Serum, Plasma or Blood by Creatinine-based formula (MDRD) (at most 20.00 mL/min/{1.73_m2}) |
|  |  | or | laboratory | UMLS:LNC:98979-8 | Glomerular filtration rate [Volume Rate/Area] in Serum, Plasma or Blood by Creatinine-based formula (CKD-EPI 2021)/1.73 sq M (at most 20.00 mL/min/{1.73_m2}) |
|  |  | or | laboratory | UMLS:LNC:50210-4 | Glomerular filtration rate [Volume Rate/Area] in Serum, Plasma or Blood by Cystatin C-based formula/1.73 sq M (at most 20.00 mL/min/{1.73_m2}) |

**Supplementary Table S4: Continued baseline characteristics before and after propensity-score matching**

|  | **Before matching** | | | **After matching** | | |
| --- | --- | --- | --- | --- | --- | --- |
|  | **SGLT2i users**  **(n= 5,045)** | **Non-users**  **(n= 370,557)** | **SMD** | **SGLT2i users**  **(n= 5,038)** | **Non-users**  **(n= 5,038)** | **SMD** |
| **Demographics** | | | | | | |
| Unknown gender | 10 (0.2%) | 350 (0.1%) | 0.018 | 10 (0.2%) | 10 (0.2%) | <0.001 |
| White | 3,007 (59.6%) | 248,888 (67.2%) | 0.157 | 3,005 (59.6%) | 3,134 (62.2%) | 0.052 |
| Black or African American | 597 (11.8%) | 39,806 (10.7%) | 0.035 | 597 (11.8%) | 516 (10.2%) | 0.051 |
| Asian* | 465 (9.2%) | 18,610 (5.0%) | 0.164 | 462 (9.2%) | 468 (9.3%) | 0.004 |
| Hispanic or latino* | 254 (5.0%) | 9,132 (2.5%) | 0.136 | 251 (5.0%) | 241 (4.8%) | 0.009 |
| Not hispanic or latino | 3,248 (64.4%) | 238,017 (64.2%) | 0.003 | 3,244 (64.4%) | 3,258 (64.7%) | 0.006 |
| Unknown ethnicity* | 811 (16.1%) | 53,646 (14.5%) | 0.044 | 809 (16.1%) | 776 (15.4%) | 0.018 |
| Other race | 133 (2.6%) | 7,058 (1.9) | 0.049 | 133 (2.6%) | 102 (2.0%) | 0.041 |
| **Comorbidities, n (%)** | | | | | | |
| Chronic obstructive pulmonary disease* | 838 (16.6%) | 52,380 (14.1%) | 0.069 | 838 (16.6%) | 868 (17.2%) | 0.016 |
| Pulmonary heart diseases and diseases of pulmonary circulation* | 830 (16.5%) | 31,511 (8.5%) | 0.242 | 826 (16.4%) | 800 (15.9%) | 0.014 |
| Diseases of arteries, arterioles and capillaries | 1,047 (20.8%) | 79,317 (21.4%) | 0.016 | 1,045 (20.7%) | 1,129 (22.4%) | 0.041 |
| Nicotine dependence* | 227 (4.5%) | 16,180 (4.4%) | 0.006 | 227 (4.5%) | 244 (4.8%) | 0.016 |
| Fibrosis and cirrhosis of liver | 86 (1.7%) | 5,582 (1.5%) | 0.016 | 86 (1.7%) | 101 (2.0%) | 0.022 |
| Benign neoplasms, except benign neuroendocrine tumors* | 257 (5.1%) | 33,932 (9.2%) | 0.158 | 257 (5.1%) | 251 (5.0%) | 0.005 |
| Neoplasms of uncertain behavior, PV and MDS* | 174 (3.4%) | 21,821 (5.9%) | 0.116 | 174 (3.5%) | 185 (3.7%) | 0.012 |
| Dementia in other diseases classified elsewhere | 122 (2.4%) | 14,727 (4.0%) | 0.089 | 122 (2.4%) | 100 (2.0%) | 0.030 |
| Other cardiac arrhythmias | 889 (17.6%) | 44,847 (12.1%) | 0.156 | 887 (17.6%) | 774 (15.4%) | 0.060 |
| Malnutrition | 270 (5.4%) | 14,838 (4.0%) | 0.064 | 270 (5.4%) | 241 (4.8%) | 0.026 |
| Do not resuscitate status | 453 (9.0%) | 19,038 (5.1%) | 0.150 | 453 (9.0%) | 420 (8.3%) | 0.023 |
| Persons encountering health services for examinations* | 1,673 (33.2%) | 160,605 (43.3%) | 0.211 | 1,672 (33.2%) | 1,683 (33.4%) | 0.005 |
| Hyperkalemia | 415 (8.2%) | 23,620 (6.4%) | 0.071 | 415 (8.2%) | 476 (9.4%) | 0.043 |
| Fluid overload* | 139 (2.8%) | 6,856 (1.9%) | 0.060 | 139 (2.8%) | 144 (2.9%) | 0.006 |
| Acidosis | 443 (8.8%) | 19,314 (5.2%) | 0.140 | 441 (8.8%) | 443 (8.8%) | 0.001 |
| Pressure ulcer | 199 (3.9%) | 10,776 (2.9%) | 0.057 | 199 (3.9%) | 202 (4.0%) | 0.003 |
| Abnormal weight loss* | 89 (1.8%) | 14,024 (3.8%) | 0.123 | 89 (1.8%) | 92 (1.8%) | 0.004 |
| Dyspnea | 1,346 (26.7%) | 74,192 (20.0%) | 0.158 | 1,342 (26.6%) | 1,314 (26.1%) | 0.013 |
| Acute respiratory failure | 767 (15.2%) | 24,073 (6.5%) | 0.283 | 765 (15.2%) | 647 (12.8%) | 0.068 |
| **Laboratory** | | | | | | |
| BMI (kg/m^2^), Mean ± SD | 28.1 ± 5.9 | 27.8 ± 5.8 | 0.044 | 28.1 ± 5.9 | 28.3 ± 6.0 | 0.044 |
| < 18.5 | 137 (2.7%) | 10,299 (2.8%) | 0.004 | 137 (2.7%) | 129 (2.6%) | 0.010 |
| 18.5 – 25 | 1,091 (21.6%) | 83,238 (22.5%) | 0.020 | 1,088 (21.6%) | 1,007 (20.0%) | 0.040 |
| 25 – 30 | 1,458 (28.9%) | 111,050 (30.0%) | 0.023 | 1,455 (28.9%) | 1,409 (28.0%) | 0.020 |
| 30 – 35 | 917 (18.2%) | 68,169 (18.4%) | 0.006 | 917 (18.2%) | 956 (19.0%) | 0.020 |
| 35 – 40 | 382 (7.6%) | 28,175 (7.6%) | 0.001 | 382 (7.6%) | 441 (8.8%) | 0.043 |
| > 40 | 185 (3.7%) | 12,208 (3.3%) | 0.020 | 185 (3.7%) | 204 (4.0%) | 0.020 |
| HbA1c (%), Mean ± SD | 7.0 ± 1.40 | 6.3 ± 1.1 | 0.541 | 7.0 ± 1.4 | 6.6 ± 1.2 | 0.253 |
| < 5 | 49 (1.0%) | 5,679 (1.5%) | 0.050 | 49 (1.0%) | 63 (1.3%) | 0.027 |
| 5 – 6 | 632 (12.5%) | 69,198 (18.7%) | 0.170 | 631 (12.5%) | 833 (16.5%) | 0.114 |
| 6 – 7 | 839 (16.6%) | 57,780 (15.6%) | 0.028 | 838 (16.6%) | 1,067 (21.2%) | 0.116 |
| 7 – 8 | 625 (12.4%) | 25,670 (6.9%) | 0.186 | 623 (12.4%) | 618 (12.3%) | 0.003 |
| 8 – 9 | 345 (6.8%) | 11,472 (3.1%) | 0.173 | 345 (6.8%) | 283 (5.6%) | 0.051 |
| > 9 | 238 (4.7%) | 7,525 (2.0%) | 0.149 | 237 (4.7%) | 159 (3.2%) | 0.080 |
| Blood pressure, systolic (mmHg)* | 130.6 ± 22.8 | 133.2 ± 20.9 | 0.119 | 130.6 ± 22.8 | 131.8 ± 22.8 | 0.055 |
| < 90 | 152 (3.0%) | 11,412 (3.1%) | 0.004 | 151 (3.0%) | 148 (2.9%) | 0.004 |
| 90 – 110 | 1,238 (24.5%) | 89,681 (24.2%) | 0.008 | 1,235 (24.5) | 1,282 (25.4%) | 0.022 |
| 110 – 120 | 1,543 (30.6%) | 131,780 (35.6%) | 0.106 | 1,539 (30.5%) | 1,580 (31.4%) | 0.018 |
| 120 – 130 | 1,939 (38.4%) | 171,889 (46.4%) | 0.161 | 1,938 (38.5%) | 1,947 (38.6%) | 0.004 |
| 130 – 140 | 1,936 (38.4%) | 180,814 (48.8%) | 0.211 | 1,935 (38.4%) | 1,997 (39.6%) | 0.025 |
| 140 – 160 | 2,102 (41.7%) | 184,724 (49.8%) | 0.165 | 2,101 (41.7%) | 2,205 (43.8%) | 0.042 |
| 160 – 180 | 1,187 (23.5%) | 111,755 (30.2%) | 0.150 | 1,187 (23.6%) | 1,232 (24.5%) | 0.021 |
| > 180 | 530 (10.5%) | 51,241 (13.8%) | 0.102 | 530 (10.5%) | 597 (11.8%) | 0.042 |

Characteristics with an asterisk were used as variables during propensity-score matching. **Abbreviations**: MDS, Myelodysplastic Syndrome; PV, Polycythemia Vera; SD, standard deviation; SMD, standardized mean difference; Std. diff., standard difference.

**Supplementary Table S5: Definitions of pre-specified outcomes**

**Main outcomes:**

| all-cause mortality (have any of the following) | | | | |
| --- | --- | --- | --- | --- |
|  | **Outcome definition** | | | |
|  | | Demographics | Deceased | Deceased |
|  | | Diagnosis | UMLS:ICD10CM:R99 | Ill-defined and unknown cause of mortality |
| major adverse kidney events (MAKE) (have any of the following) | | | | |
|  | **Outcome definition** | | | |
|  | | Demographics | Deceased | Deceased |
|  | | Diagnosis | UMLS:ICD10CM:R99 | Ill-defined and unknown cause of mortality |
|  | | Procedure | UMLS:CPT:1012740 | Dialysis Services and Procedures |
|  | | Diagnosis | UMLS:ICD10CM:N18.5 | Chronic kidney disease, stage 5 |
|  | | Diagnosis | UMLS:ICD10CM:N18.6 | End stage renal disease |
|  | | Diagnosis | UMLS:ICD10CM:Z99.2 | Dependence on renal dialysis |
|  | | Procedure | UMLS:ICD10PCS:5A1D | Physiological Systems / Performance / Urinary |
|  | | Procedure | UMLS:SNOMED:302497006 | Hemodialysis |
| major adverse cardiovascular events (MACE) (have any of the following) | | | | |
|  | **Outcome definition** | | | |
|  | | Demographics | Deceased | Deceased |
|  | | Diagnosis | UMLS:ICD10CM:R99 | Ill-defined and unknown cause of mortality |
|  | | Diagnosis | UMLS:ICD10CM:I60 | Nontraumatic subarachnoid hemorrhage |
|  | | Diagnosis | UMLS:ICD10CM:I61 | Nontraumatic intracerebral hemorrhage |
|  | | Diagnosis | UMLS:ICD10CM:I63 | Cerebral infarction |
|  | | Diagnosis | UMLS:ICD10CM:I21 | Acute myocardial infarction |
|  | | Diagnosis | UMLS:ICD10CM:I46 | Cardiac arrest |

**Specific kidney outcomes**

| acute kidney injury (AKI) (have any of the following) | | | | |
| --- | --- | --- | --- | --- |
|  | **Outcome definition** | | | |
|  | | Diagnosis | UMLS:ICD10CM:N17 | Acute kidney failure |
| Incident Dialysis (have any of the following) | | | | |
|  | **Outcome definition** | | | |
|  | | Procedure | UMLS:CPT:1012740 | Dialysis Services and Procedures |
|  | | Diagnosis | UMLS:ICD10CM:Z99.2 | Dependence on renal dialysis |
|  | | Procedure | UMLS:ICD10PCS:5A1D | Physiological Systems / Performance / Urinary |
|  | | Procedure | UMLS:SNOMED:302497006 | Hemodialysis |
| end-stage renal disease (ESRD) (have any of the following) | | | | |
|  | **Outcome definition** | | | |
|  | | Diagnosis | UMLS:ICD10CM:N18.6 | End stage renal disease |
|  | | Diagnosis | UMLS:ICD10CM:N18.5 | Chronic kidney disease, stage 5 |

**Specific cardiovascular outcomes**

| acute myocardial infarction (AMI) | | | | |
| --- | --- | --- | --- | --- |
|  | **Outcome definition** | | | |
|  | | Diagnosis | UMLS:ICD10CM:I21 | Acute myocardial infarction |
| Cardiac arrest | | | | |
|  | **Outcome definition** | | | |
|  | | Diagnosis | UMLS:ICD10CM:I46 | Cardiac arrest |
| Acute heart failure (have any of the following) | | | | |
|  | **Outcome definition** | | | |
|  | | Diagnosis | UMLS:ICD10CM:I50.41 | Acute combined systolic (congestive) and diastolic (congestive) heart failure |
|  | | Diagnosis | UMLS:ICD10CM:I50.43 | Acute on chronic combined systolic (congestive) and diastolic (congestive) heart failure |
|  | | Diagnosis | UMLS:ICD10CM:I50.813 | Acute on chronic right heart failure |
|  | | Diagnosis | UMLS:ICD10CM:I50.23 | Acute on chronic systolic (congestive) heart failure |
|  | | Diagnosis | UMLS:ICD10CM:I50.33 | Acute on chronic diastolic (congestive) heart failure |
|  | | Diagnosis | UMLS:ICD10CM:I50.811 | Acute right heart failure |
|  | | Diagnosis | UMLS:ICD10CM:I50.31 | Acute diastolic (congestive) heart failure |
|  | | Diagnosis | UMLS:ICD10CM:I50.21 | Acute systolic (congestive) heart failure |
| Cerebral infarction | | | | |
| Outcome definition | | | | |
|  | | Diagnosis | UMLS:ICD10CM:I63 | Cerebral infarction |
| Cerebral bleeding (have any of the following | | | | |
| Outcome definition | | | | |
|  | | Diagnosis | UMLS:ICD10CM:I60 | Nontraumatic subarachnoid hemorrhage |
|  | | Diagnosis | UMLS:ICD10CM:I61 | Nontraumatic intracerebral hemorrhage |
| combined stroke (have any of the following) | | | | |
| Outcome definition | | | | |
|  | | Diagnosis | UMLS:ICD10CM:I60 | Nontraumatic subarachnoid hemorrhage |
|  | | Diagnosis | UMLS:ICD10CM:I61 | Nontraumatic intracerebral hemorrhage |
|  | | Diagnosis | UMLS:ICD10CM:I63 | Cerebral infarction |

**Adverse events:**

| Urinary tract infection (UTI) (have any of the following) | | | | |
| --- | --- | --- | --- | --- |
|  | **Outcome definition** | | | |
|  | | Diagnosis | UMLS:ICD10CM:N39.0 | Urinary tract infection, site not specified |
|  | | Diagnosis | UMLS:ICD10CM:N30 | Cystitis |
|  | | Diagnosis | UMLS:ICD10CM:N10 | Acute pyelonephritis |
| Genitourinary Candidiasis (have any of the following) | | | | |
|  | **Outcome definition** | | | |
|  | | Diagnosis | UMLS:ICD10CM:B37.3 | Candidiasis of vulva and vagina |
|  | | Diagnosis | UMLS:ICD10CM:B37.4 | Candidiasis of other urogenital sites |
| Osteoporotic fracture | | | | |
|  | **Outcome definition** | | | |
|  | | Diagnosis | UMLS:ICD10CM:M80 | Osteoporosis with current pathological fracture |

| Ketoacidosis | | | |
| --- | --- | --- | --- |
| Outcome definition | | | |
|  | Diagnosis | UMLS:ICD10CM:E11.1 | Type 2 diabetes mellitus with ketoacidosis |

**Unrelated events:**

| hematologic neoplasms | | | | |
| --- | --- | --- | --- | --- |
|  | **Outcome definition** | | | |
|  | | Diagnosis | UMLS:ICD10CM:C81-C96 | Malignant neoplasms of lymphoid, hematopoietic and related tissue |
| Gastritis | | | | |
|  | **Outcome definition** | | | |
|  | | Diagnosis | UMLS:ICD10CM:K29 | Gastritis and duodenitis |

**Supplementary Table S6: Landmark analyses with the follow-up period starting at different time points after the index event**

**S6A: follow-up period 3 months until 2 years after the index event**

| **Outcome** | **No. of patients with outcome** | | **aHR** | **95% CI** | **p** |
| --- | --- | --- | --- | --- | --- |
|  | **SGLT2i** | **no-SGLT2i** |  |  |  |
| All-cause mortality | 502 | 681 | 0.787 | 0.701-0.883 | <0.0001 |
| MAKE | 577 | 813 | 0.745 | 0.669-0.829 | <0.0001 |
| MACE | 959 | 1,056 | 0.983 | 0.901-1.073 | 0.6996 |

**S6B: follow-up period 6 months until 2 years after the index event**

| **Outcome** | **No. of patients with outcome** | | **aHR** | **95% CI** | **p** |
| --- | --- | --- | --- | --- | --- |
|  | **SGLT2i** | **no-SGLT2i** |  |  |  |
| All-cause mortality | 390 | 527 | 0.797 | 0.699-0.908 | 0.0006 |
| MAKE | 460 | 645 | 0.756 | 0.671-0.852 | <0.0001 |
| MACE | 781 | 852 | 0.996 | 0.904-1.098 | 0.9354 |

**Abbreviations:** MAKE, major adverse kidney events; MACE, major adverse cardiovascular events; 95% CI, 95% confidence interval.

**Supplementary Table S7: Sensitivity analyses at different time points**

|  | *3 months* | *6 months* | *1 year* | *1.5 years* | *3 years* |
| --- | --- | --- | --- | --- | --- |
| **Mortality** | 0.882  (0.759-1.024) | 0.843  (0.742-0.958) | 0.834  (0.749-0.928) | 0.835  (0.757-0.920) | 0.835  (0.766-0.909) |
| **MAKE** | 0.752  (0.653-0.865) | 0.733  (0.650-0.827) | 0.769  (0.695-0.850) | 0.777  (0.709-0.851) | 0.801  (0.739-0.869) |
| **MACE** | 1.120  (1.008-1.245) | 1.087  (0.990-1.193) | 1.027  (0.948-1.114) | 1.027  (0.952-1.107) | 1.013  (0.946-1.084) |

Adjusted hazard ratios and 95% confidence intervals are shown.
**Abbreviations:** MAKE, major adverse kidney events; MACE, major adverse cardiovascular events;

**Supplementary Table S8: The RECORD statement – checklist of items, extended from the STROBE statement**

|  | **Item No.** | **STROBE items** | **Location in manuscript where items are reported** | **RECORD items** | **Location in manuscript where items are reported** |
| --- | --- | --- | --- | --- | --- |
| **Title and abstract** | | | | | |
|  | 1 | (a) Indicate the study’s design with a commonly used term in the title or the abstract (b) Provide in the abstract an informative and balanced summary of what was done and what was found | (a) p. 1  (b) p. 2 | RECORD 1.1: The type of data used should be specified in the title or abstract. When possible, the name of the databases used should be included.  RECORD 1.2: If applicable, the geographic region and timeframe within which the study took place should be reported in the title or abstract.  RECORD 1.3: If linkage between databases was conducted for the study, this should be clearly stated in the title or abstract. | 1.1: p. 1  1.2: p. 2+5  1.3: n.a. |
| **Introduction** | | | | | |
| Background rationale | 2 | Explain the scientific background and rationale for the investigation being reported | pp. 2-3 |  |  |
| Objectives | 3 | State specific objectives, including any prespecified hypotheses | p. 3 |  |  |
| **Methods** | | | | | |
| Study Design | 4 | Present key elements of study design early in the paper | pp. 5-6 |  |  |
| Setting | 5 | Describe the setting, locations, and relevant dates, including periods of recruitment, exposure, follow-up, and data collection | pp. 5-6 |  |  |
| Participants | 6 | *(a) Cohort study* - Give the eligibility criteria, and the sources and methods of selection of participants. Describe methods of follow-up  *Case-control study* - Give the eligibility criteria, and the sources and methods of case ascertainment and control selection. Give the rationale for the choice of cases and controls  *Cross-sectional study* - Give the eligibility criteria, and the sources and methods of selection of participants  *(b) Cohort study* - For matched studies, give matching criteria and number of exposed and unexposed  *Case-control study* - For matched studies, give matching criteria and the number of controls per case | (a) pp. 5-6  (b) pp. 17-20, suppl. pp. 8-9 | RECORD 6.1: The methods of study population selection (such as codes or algorithms used to identify subjects) should be listed in detail. If this is not possible, an explanation should be provided.  RECORD 6.2: Any validation studies of the codes or algorithms used to select the population should be referenced. If validation was conducted for this study and not published elsewhere, detailed methods and results should be provided.  RECORD 6.3: If the study involved linkage of databases, consider use of a flow diagram or other graphical display to demonstrate the data linkage process, including the number of individuals with linked data at each stage. | 6.1: suppl. pp. 6-7  6.2: detailed methods provided, additional reference: 22 (Biswas et al.)  6.3: n.a. |
| Variables | 7 | Clearly define all outcomes, exposures, predictors, potential confounders, and effect modifiers. Give diagnostic criteria, if applicable. | pp. 17-18  suppl. pp. 4-5, 8-9, 10-11  discussion on pp. 11-12 | RECORD 7.1: A complete list of codes and algorithms used to classify exposures, outcomes, confounders, and effect modifiers should be provided. If these cannot be reported, an explanation should be provided. | suppl. pp. 4-7, 10-11 |
| Data sources/ measurement | 8 | For each variable of interest, give sources of data and details of methods of assessment (measurement).  Describe comparability of assessment methods if there is more than one group | see above, pp. 5-6 |  |  |
| Bias | 9 | Describe any efforts to address potential sources of bias | p. 5, discussion on pp. 11-12 |  |  |
| Study size | 10 | Explain how the study size was arrived at | pp. 5-7, 20 (Fig. 1) |  |  |
| Quantitative variables | 11 | Explain how quantitative variables were handled in the analyses. If applicable, describe which groupings were chosen, and why | suppl. p. 9 |  |  |
| Statistical methods | 12 | (a) Describe all statistical methods, including those used to control for confounding  (b) Describe any methods used to examine subgroups and interactions  (c) Explain how missing data were addressed  (d) *Cohort study* - If applicable, explain how loss to follow-up was addressed  *Case-control study* - If applicable, explain how matching of cases and controls was addressed  *Cross-sectional study* - If applicable, describe analytical methods taking account of sampling strategy  (e) Describe any sensitivity analyses | (a) pp. 5-6  (b) p. 6  (c) p. 7, p. 14  (d) p. 7, p. 14  (e) pp. 7, 8-9 |  |  |
| Data access and cleaning methods |  | .. |  | RECORD 12.1: Authors should describe the extent to which the investigators had access to the database population used to create the study population.  RECORD 12.2: Authors should provide information on the data cleaning methods used in the study. | 12.1: p. 14  12.2: n.a. |
| Linkage |  | .. |  | RECORD 12.3: State whether the study included person-level, institutional-level, or other data linkage across two or more databases. The methods of linkage and methods of linkage quality evaluation should be provided. | 12.3: p.6 |
| **Results** | | | | | |
| Participants | 13 | (a) Report the numbers of individuals at each stage of the study (*e.g.*, numbers potentially eligible, examined for eligibility, confirmed eligible, included in the study, completing follow-up, and analysed)  (b) Give reasons for non-participation at each stage.  (c) Consider use of a flow diagram | Fig. 1 | RECORD 13.1: Describe in detail the selection of the persons included in the study (*i.e.,* study population selection) including filtering based on data quality, data availability and linkage. The selection of included persons can be described in the text and/or by means of the study flow diagram. | Fig. 1, Suppl. Tables S2+S3 |
| Descriptive data | 14 | (a) Give characteristics of study participants (*e.g.*, demographic, clinical, social) and information on exposures and potential confounders  (b) Indicate the number of participants with missing data for each variable of interest  (c) *Cohort study* - summarise follow-up time (*e.g.*, average and total amount) | Table 1 and Suppl. Table S4 |  |  |
| Outcome data | 15 | *Cohort study* - Report numbers of outcome events or summary measures over time  *Case-control study* - Report numbers in each exposure category, or summary measures of exposure  *Cross-sectional study* - Report numbers of outcome events or summary measures | pp. 2, 8-9, Table 2, Fig. 2-5, Suppl. Tables S6+S7 |  |  |
| Main results | 16 | (a) Give unadjusted estimates and, if applicable, confounder-adjusted estimates and their precision (e.g., 95% confidence interval). Make clear which confounders were adjusted for and why they were included  (b) Report category boundaries when continuous variables were categorized  (c) If relevant, consider translating estimates of relative risk into absolute risk for a meaningful time period | (a) pp. 2, 8-9, Table 2, Fig. 2-5, Suppl. Tables S6+S7, Table 1 + Suppl. Table S4, p. 6  (b) Table 1 + Suppl. Table S4  (c) n.a. |  |  |
| Other analyses | 17 | Report other analyses done—e.g., analyses of subgroups and interactions, and sensitivity analyses | Fig. 2+3, Suppl. Fig. S2, Suppl. Tables S6+S7 |  |  |
| **Discussion** | | | | | |
| Key results | 18 | Summarise key results with reference to study objectives | pp. 10, 13 |  |  |
| Limitations | 19 | Discuss limitations of the study, taking into account sources of potential bias or imprecision. Discuss both direction and magnitude of any potential bias | pp.12-13 | RECORD 19.1: Discuss the implications of using data that were not created or collected to answer the specific research question(s). Include discussion of misclassification bias, unmeasured confounding, missing data, and changing eligibility over time, as they pertain to the study being reported. | pp. 12-13 |
| Interpretation | 20 | Give a cautious overall interpretation of results considering objectives, limitations, multiplicity of analyses, results from similar studies, and other relevant evidence | pp. 10-13 |  |  |
| Generalisability | 21 | Discuss the generalisability (external validity) of the study results | pp. 12-13 |  |  |
| **Other Information** | | | | | |
| Funding | 22 | Give the source of funding and the role of the funders for the present study and, if applicable, for the original study on which the present article is based | p. 14 |  |  |
| Accessibility of protocol, raw data, and programming code |  | .. |  | RECORD 22.1: Authors should provide information on how to access any supplemental information such as the study protocol, raw data, or programming code. | p. 14 |

*Reference: Benchimol EI, Smeeth L, Guttmann A, Harron K, Moher D, Petersen I, Sørensen HT, von Elm E, Langan SM, the RECORD Working Committee. The REporting of studies Conducted using Observational Routinely-collected health Data (RECORD) Statement. *PLoS Medicine* 2015; https://doi.org/10.1371/journal.pmed.1001885.

*Checklist is protected under Creative Commons Attribution ([CC BY](http://creativecommons.org/licenses/by/4.0/)) license.
